# The accuracy of urine-based mycobacterial antigens to detect childhood tuberculosis using an ultrasensitive immunoassay

**DOI:** 10.64898/2026.08.28.26361530

**Authors:** Esin Nkereuwem, Salvia Misaghian, Devan Jaganath, Roger I. Calderon, Juaneta Luiz, Mandar Paradkar, Peter Wambi, Robert Castro, Rutuja Nerurkar, Mingyue Wang, Jacob Wohlstadter, Molly F. Franke, Beate Kampmann, Aarti Kinikar, Heather J. Zar, Mark Segal, Midori Kato-Maeda, Jeffrey M. Collins, Danielle Swaney, Adithya Cattamanchi, Joel D. Ernst, Eric Wobudeya, George B. Sigal, the COMBO study

## Abstract

**Background:** Urine-based testing offers a promising non-sputum approach for diagnosing paediatric tuberculosis. However, the currently available lipoarabinomannan (LAM) assay shows limited sensitivity in children and is primarily indicated for those living with HIV. Co-detection of LAM with *Mycobacterium tuberculosis* (*Mtb*) proteins in urine could provide complementary pathogen-derived biomarkers that improve diagnostic performance.

**Methods:** We developed an ultrasensitive multiplex electrochemiluminescence (ECL) immunoassay to measure Ag85B, CFP-10, ESAT-6, MPT32, and MPT64 in urine. We determined the analytical limits of detection and evaluated the diagnostic performance of individual proteins and LAM using urine samples from children with Confirmed, Unconfirmed, and Unlikely pulmonary tuberculosis enrolled across five high-burden countries (The Gambia, India, Peru, South Africa, and Uganda). Performance was assessed overall, by HIV and nutritional status, and across biomarker combinations.

**Findings:** Urine samples from 630 children were analysed (median age was 4 years [IQR 2-8]; 44% female, 15% living with HIV, 19% underweight, 24% with Confirmed tuberculosis). The ECL assay achieved femtomolar limits of detection (1·5 to 4·0 fM). The sensitivity and specificity of individual *Mtb* proteins were 12-33% and 98-100%, respectively. Ag85B had the highest sensitivity (33%, 95% CI 26-41) for Confirmed tuberculosis and was similar to LAM. A four-antigen signature (Ag85B, MPT64, MPT32, LAM) was 50% sensitive (95% CI 42-58) and 94% specific (95% CI 90-96), and was significantly more sensitive than LAM alone, in particular among those without HIV. An additional sixteen (10%) of children with Unconfirmed TB had at least one *Mtb* protein or LAM detected.

**Interpretation:** Multiple *Mtb* proteins are detectable in paediatric urine with high specificity, and multi-antigen signatures can augment sensitivity versus LAM alone. These findings demonstrate the potential of multi-antigen urine detection for childhood TB and define analytical targets for the development of future point-of-care diagnostics.

**Funding:** National Institutes of Health.

**RESEARCH IN CONTEXT:** *Evidence before this study:* We examined the literature for peer-reviewed research articles on the accuracy of biomarker-based urine tests for pulmonary tuberculosis in children <15 years old. We used PubMed and Google Scholar, with the search terms “child”, “tuberculosis”, “urine”, and “diagnosis” regardless of language from July 2016 to July 2026. We excluded articles on host-based markers and extrapulmonary tuberculosis. Molecular urine assays, including Xpert MTB/RIF, have limited sensitivity to detect childhood pulmonary tuberculosis. Most of the research on urine-based diagnostics has focused on detection of lipoarabinomannan (LAM), which has had variable sensitivity and specificity in children against a microbiological reference standard. Accuracy is higher in those with HIV, and current guidelines only recommend LAM for adults and children with HIV.

*Added value of this study:* We developed an ultrasensitive multiplex immunoassay to detect and measure *Mtb-*specific proteins in urine samples from children with presumptive tuberculosis in five high-burden countries. We found that *Mtb* proteins could be detected in paediatric urine samples with high specificity, and Ag85B had similar sensitivity as LAM. A four-marker panel (Ag85B, MPT32, MPT64, and LAM) improved sensitivity over LAM alone without a significant loss of specificity, in particular among children without HIV.

*Implications of all the available evidence:* Multi-antigen urine tests can improve sensitivity over single marker assays, and have the potential to provide non-sputum, point-of-care tuberculosis detection in children regardless of HIV status.

## INTRODUCTION

Tuberculosis, caused by *Mycobacterium tuberculosis* (*Mtb*), remains a leading cause of death from an infectious disease worldwide. Notably, children and young adolescents under 15 years of age bear a disproportionate burden of tuberculosis mortality, accounting for 12% of incident cases but 15% of tuberculosis-related deaths globally.^1^ Diagnosis of pulmonary tuberculosis in children remains particularly challenging because respiratory specimens are difficult to obtain and typically contain low bacillary loads, limiting the sensitivity of standard culture, smear microscopy, and molecular testing.^2^ As a result, a substantial proportion of childhood tuberculosis cases remain undetected or unreported, highlighting the need for alternative, non-sputum-based diagnostic approaches.^1^

Urine represents an attractive diagnostic specimen because it can be obtained non-invasively, safely, and repeatedly, avoiding the technical and biosafety challenges of paediatric sputum collection. Consequently, detection of *Mtb* lipoarabinomannan (LAM) antigen has been developed as a non-sputum diagnostic approach.^3^ However, its performance remains variable and suboptimal. Reported sensitivity ranges from 13% to 51% in adults,^4^ and 28% to 73% in children,^5^ with substantially lower sensitivity in HIV-negative individuals. Lateral flow LAM assays perform best in people living with HIV and advanced immunosuppression, and the WHO currently recommends their use primarily in adults and children with HIV.^6^ However, the diagnostic yield in HIV-negative children remains insufficient. Specificity may also be reduced in young children, particularly with bagged urine collection, possibly due to cross-reactivity with non-tuberculous mycobacteria and other bacteria.^7^ Even next-generation LAM assays have generally failed to meet the WHO target product profile benchmarks for non-sputum point-of-care diagnostics (minimum ≥65% sensitivity and ≥98% specificity).^8^

These limitations suggest that LAM alone may be insufficient for reliable diagnosis of tuberculosis in children. Several *Mtb* proteins have been investigated as diagnostic biomarkers, including Ag85B, CFP-10, ESAT-6, MPT32, and MPT64.^9,10^ Importantly, some of these proteins have also been detected in urine from individuals with active tuberculosis, supporting the feasibility of urine-based antigen detection.^9,10^ These findings suggest that combining multiple *Mtb* antigens with LAM could enhance diagnostic sensitivity compared with single-analyte approaches without substantially compromising specificity.

In this study, we evaluated five *Mtb* proteins, namely Ag85B, CFP-10, ESAT-6, MPT32, and MPT64, that are either secreted at high levels or have been previously detected in urine.^9,10^ Building on earlier work showing urinary detection of ESAT-6,^11,12^ we developed a multiplex ultrasensitive electrochemiluminescence (ECL) immunoassay on the S-PLEX^®^ platform (Meso Scale Discovery (MSD), Rockville, Maryland). We evaluated the diagnostic performance of this assay in a multi-country cohort of children presenting with symptoms suggestive of tuberculosis. For benchmarking, we compared its performance with a previously developed high-sensitivity urinary LAM assay,^12^ which has also been used to evaluate new *Mtb* antigen-based tests.^13^

## METHODS

### Clinical Samples

We analysed stored urine samples from adults and children as part of the Childhood “Omics” and *Mycobacterium tuberculosis*-derived BiOsignatures (COMBO) study, a multi-country initiative investigating host and pathogen biomarkers for childhood tuberculosis.^14^ Paediatric participants had previously been enrolled in prospective diagnostic cohort studies evaluating children younger than 15 years presenting with symptoms suggestive of pulmonary tuberculosis in The Gambia, Peru, South Africa, and Uganda. Urine samples were collected at enrolment and stored at −80°C at the study sites.

#### Adult Sample Set

Adult urine samples were used for initial assay threshold identification. Sixty de-identified urine samples were selected from FIND’s biobank, from South-East Asia (Cambodia, n = 6, and Vietnam, n = 18), South America (Peru, n = 13) and Africa (South Africa, n = 23), who presented with clinical symptoms of tuberculosis and had not yet started tuberculosis treatment.^12,15^ The donors were 37% female with a median age of 38 years (IQR 32 to 48).

Tuberculosis-positive individuals were positive by both sputum smear and culture. Tuberculosis- negative individuals were sputum smear and culture negative, exhibited symptom resolution in the absence of tuberculosis treatment, and had negative sputum culture at a 2-month follow-up visit. Approval by local ethics committees and informed patient consent were obtained. The samples were selected to include an equal number (15) of each possible combination of tuberculosis and HIV status.

#### Paediatric Sample Set

The paediatric samples analysed in this study were collected between December 2010, and March 2021 from five countries: The Gambia, India, Peru, South Africa, and Uganda. Study sites included the Medical Research Council (MRC) Unit The Gambia at the London School of Hygiene & Tropical Medicine, Fajara, The Gambia; Byramjee Jeejeebhoy Medical College, Pune, India; Socios en Salud, Lima, Peru; University of Cape Town and Dora Nginza Hospital, Gqeberha, South Africa; and Mulago National Referral Hospital in Kampala, Uganda.

Children younger than 15 years were eligible if they had at least one respiratory specimen collected for microbiological testing. Clinical procedures are described in the **Supplemental Methods**. All children were classified into three diagnostic categories by expert clinicians at each site, following NIH consensus definitions: Confirmed tuberculosis, Unconfirmed tuberculosis, and Unlikely tuberculosis.^16^ Confirmed tuberculosis required microbiological confirmation of *Mtb*, defined as Xpert or culture positive from a respiratory sample. Unconfirmed tuberculosis referred to children with negative microbiological tests for tuberculosis who, based on clinical or radiographic evidence, were started on tuberculosis treatment and showed improvement. Unlikely tuberculosis described symptomatic children who had negative microbiological tests, no clinical evidence of tuberculosis disease, and improved without tuberculosis treatment. We purposely selected stored urine samples to have approximately a 1:1:2 ratio of Confirmed:Unconfirmed:Unlikely tuberculosis.

### *Mtb* Protein 5-Plex Assay

The protein biomarkers Ag85B, CFP-10, ESAT-6, MPT32, and MPT64 were measured using a prototype multiplexed immunoassay kit using the MSD^®^ S-PLEX assay format, which employs ECL detection and a proprietary signal enhancement step to increase assay sensitivity.^17^ Assays were run in MSD S-PLEX 96-Well SECTOR plates with 10 unique “spots” or array elements, each with a different binding reagent. Assay reagents, generation of monoclonal antibodies and the assay protocol are described in the **Supplemental Methods**.

### LAM Assay

LAM was measured using a non-enhanced prototype ECL immunoassay that has been described in detail previously.^12^ The multiplexed format uses a single SULFO-TAG^TM^ labeled LAM detection antibody (A194-01) and an array of two capture antibodies (S4-20 and FIND28). Because the S4- 20 capture antibody, which targets a unique 5-methylthio-D-xylofuranose (MTX) on *Mtb* LAM, was previously determined to provide better sensitivity and specificity for urine samples,^12^ only the data generated with the S4-20 capture antibody is reported here.

### Sample Testing

Urine samples were run using the multiplexed *Mtb* Protein 5-Plex and LAM assays in duplicate without dilution or pre-treatment. Each plate run also included eight calibration samples run in duplicate, generated by 1:4 serial dilutions of a calibration standard containing a mixture of the recombinant analytes (protein assay) or purified LAM (LAM assay). Each plate run additionally included three control samples (high positive, low positive and negative) run in duplicate. Calculation of the limit of detection (LOD), top of the curve (TOC), and lower limit of quantitation (LLOQ) are described in the Supplemental Methods.

### Data and Statistical Analysis

With the adult sample measurements, we first generated a receiver operating characteristic (ROC) curve for each antigen and selected an optimal cut-off value to maximise sensitivity + 2 times the specificity. This would bias towards the higher specificity needed for a tuberculosis diagnostic (≥98%). We then calculated the sensitivity and specificity with 95% confidence intervals (CIs) at this threshold with the paediatric samples, in order to apply an independently derived cut- off value. In addition, we generated an ROC curve for each antigen with the paediatric data, and re-calculated sensitivity and specificity with a child-specific cut-off value using the same goal to maximise specificity. We measured accuracy overall, by country, and for key subgroups of children living with and without HIV, who were underweight (defined as a WHO weight-for-age z-score <-2), and those less than 5 years old. We compared the accuracy across antigens using McNemar’s test with significance in difference defined as a p-value < 0·05.

We also determined the change in accuracy if antigens were combined. We evaluated every combination panel from 1-6 antigens (n=63), and considered the panel to be positive if any one antigen was positive according to their individual cut-off. We then ranked them according to highest sensitivity. We performed this overall and stratified by HIV and underweight status. Estimates of the absolute difference in sensitivity and specificity were compared to the LAM assay alone using McNemar’s test.

Data and statistical analyses were carried out using R statistical v4.5.1, with the following packages: *prop.test, pROC,* and *ggplot2*.

## RESULTS

### Paediatric Cohort Characteristics

**Table 1** summarises the characteristics of the paediatric sample set. The overall median age was 4 years (IQR 2-8), with 44% female, 15% living with HIV and 19% underweight. Forty-nine percent were diagnosed with tuberculosis disease, of which half had Confirmed tuberculosis (n=153). Some characteristics varied across sites; notably, in South Africa, age was lower (median 2 years), HIV prevalence higher (47%) and a higher proportion had Confirmed tuberculosis (39%). Underweight status also ranged from 0% in Peru to 31% in the Gambia.

**Table 1.**
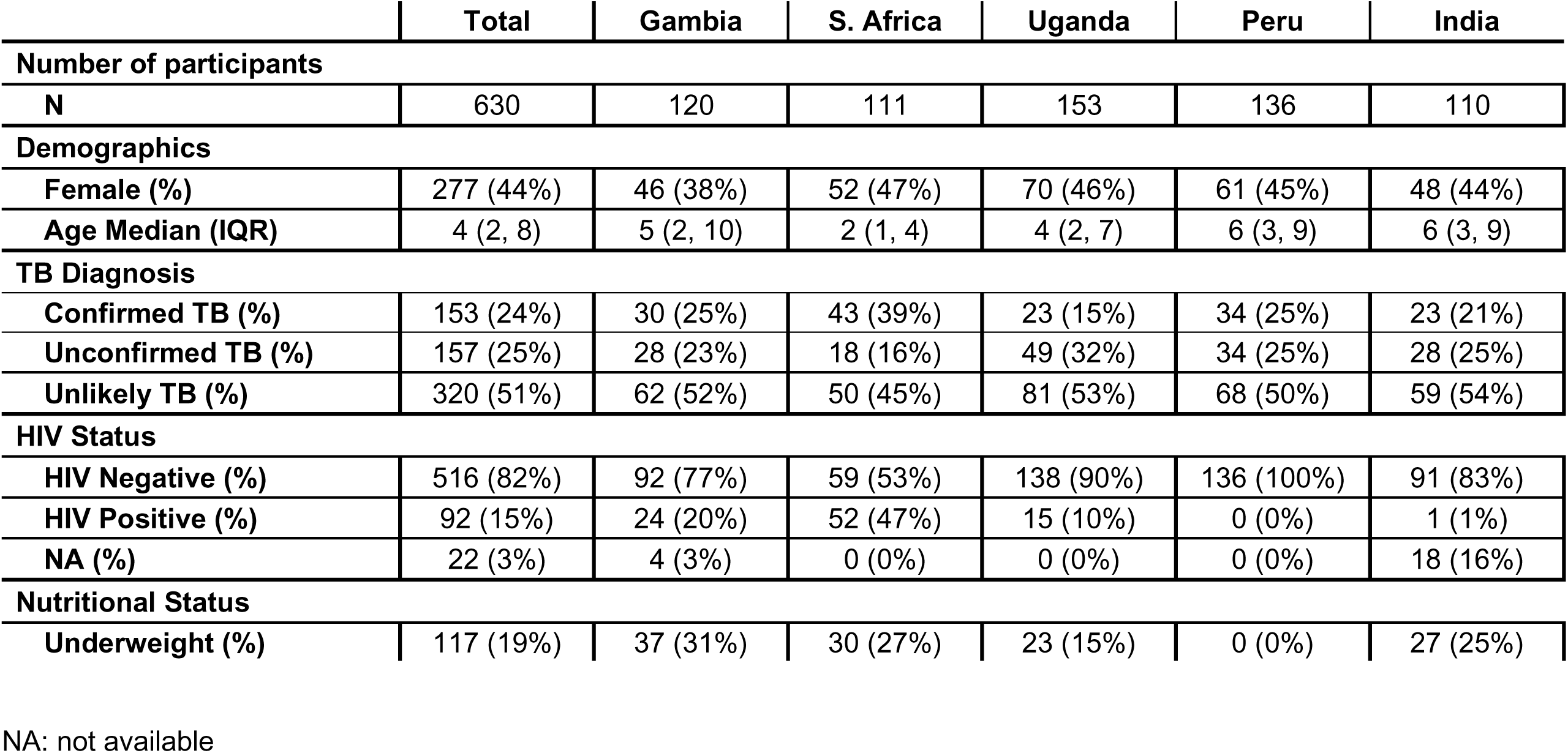
Demographic and clinical characteristics of the paediatric cohort.

### Assay Quantitation Performance

The LOD, LLOQ and TOC for the *Mtb* Protein 5-Plex and LAM assays are tabulated in **Table 2**, and annotated on typical assay calibration curves in **Supplementary Figure 1**. The LODs for the protein assays were all low fM, ranging from 1·5 fM (51 fg/mL) for Ag85B to 4·0 fM (44 fg/mL) for CFP-10. The assays provided good reproducibility across the runs. The coefficients of variation (CVs) for the measured concentrations of the low and high positive controls over all the runs were less than 25% for all assays. The median CVs of duplicate urine sample measurements for samples above the LLOQ ranged from 4% for Ag85B to 17% for MPT64.

**Table 2.**
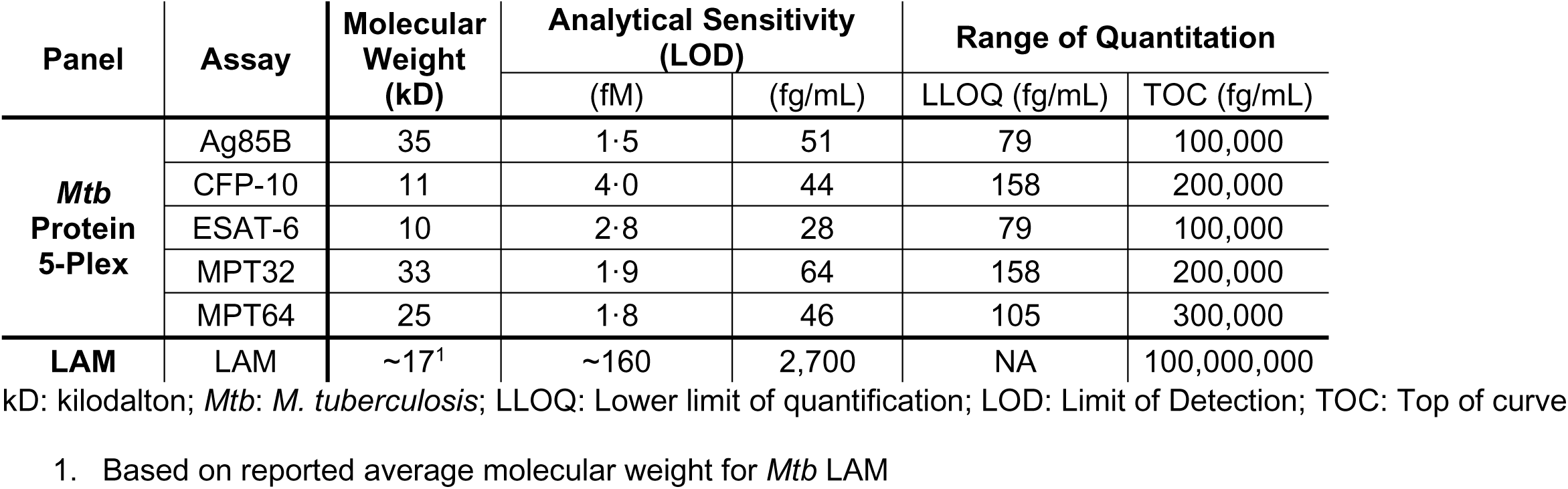
Analytical performance of *Mtb* Protein 5-Plex and LAM Assays.

### Setting Preliminary Assay Thresholds with an Adult Sample Set

**Supplementary Figure 2** shows the measured *Mtb* biomarker levels for the adult tuberculosis cohort. **Supplementary Figure 3a** shows ROC curves for each of the assays, characterizing the ability of the assays to classify samples as TB positive or negative. **Supplementary Table 1** lists the selected cut-off values and classification performance for each assay.

### Measuring Assay Performance with the Paediatric Sample Set

**Figure 1** shows the measured *Mtb* biomarker levels for the paediatric tuberculosis cohort, comparing the observed levels for the Confirmed tuberculosis, Unconfirmed tuberculosis and Unlikely tuberculosis groups. **Supplementary Table 2** provides the classification performance for each assay using the preliminary cut-off values optimised for the adult cohort. The assays maintained high specificity for classifying the Unlikely tuberculosis cases, and only one assay had a measured specificity less than 90% (MPT64, specificity 82%). Specificity values for the other assays ranged from 92% to 98%. **Figure 1** shows that all the tuberculosis biomarkers were observed at high levels in at least some Confirmed tuberculosis case samples. Sensitivity for classifying Confirmed tuberculosis samples ranged from 14% to 39%, with three assays providing sensitivity values greater than 30%: Ag85B (39%), MPT64 (33%) and LAM (35%). **Figure 1** also shows a lower frequency of positive samples and lower biomarker concentrations for the Unconfirmed tuberculosis group. As a result, expanding the case definition to include the Unconfirmed tuberculosis group lowered the observed sensitivities across all assays (Range 9- 23%).

**Figure 1.**
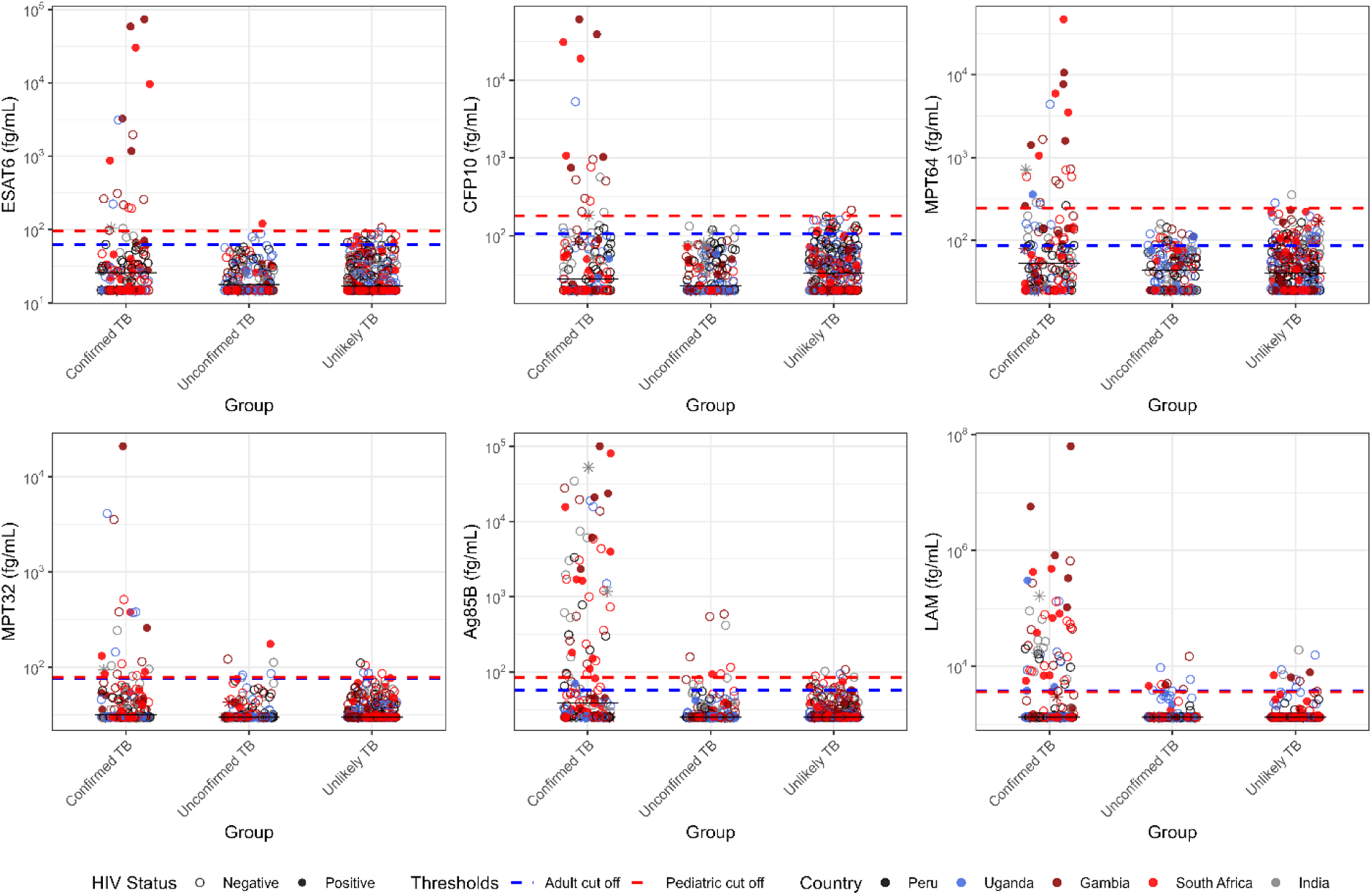
Measured concentrations of *Mtb* biomarkers in the paediatric tuberculosis urine sample set. Points are colored to indicate country, and shaped based on HIV status. The dashed horizontal lines indicate the preliminary cut-off value based on ROC curve analysis of the adult samples (blue) and the revised cut-off value (red) after optimising for the paediatric sample set.

We analysed assay performance by subgroup and geographical region (**Supplemental Table 2**). The sensitivity of all assays was higher for children living with HIV, and for underweight children. In contrast, age (above or below 5 years) did not appear to affect assay sensitivity. Sensitivity for all the assays tended to be lower for children in Uganda and Peru.

### Re-optimising Assay Thresholds for the Paediatric Sample Set

ROC analysis was repeated for the paediatric sample set to distinguish Confirmed tuberculosis from Unlikely tuberculosis (**Supplementary Figure 3b**). **Table 3** shows the adjusted cut-off values generated using the paediatric ROC curves, and the associated specificity and sensitivity values. For the *Mtb* protein assays, the revised cut-off values were generally higher than the preliminary values, providing a boost in specificity (Ranging from 98% to 100%, overall and among subgroups), but at a cost in sensitivity (Ranging from 12% to 33% for Confirmed tuberculosis cases). LAM specificity was 97% (95% CI 94-98).

**Table 3.** Classification performance of MTB Protein 5-Plex and LAM Assays with the paediatric cohort using adjusted child- specific cut-off values.

|  |  | Mtb Protein 5-Plex |  |  |  |  |  |
| --- | --- | --- | --- | --- | --- | --- | --- |
|  | N | Ag85B | CFP10 | ESAT6 | MPT32 | MPT64 | LAM |
| **Cut-Off (fg/mL)<sup>1</sup>** |  | 85 | 181 | 96 | 79 | 246 | 3656 |
| **AUC** |  | 0.72 | 0.52 | 0.58 | 0.6 | 0.62 | 0.7 |
| **Specificity, % (95% CI)** |  |  |  |  |  |  |  |
| **Unlikely TB** | 320 | 99% (97%, 100%) | 100% (98%, 100%) | 100% (98%, 100%) | 98% (96%, 99%) | 99% (98%, 100%) | 97% (94%, 98%) |
| **HIV Positive** | 42 | 100% (90%, 100%) | 100% (90%, 100%) | 100% (90%, 100%) | 100% (90%, 100%) | 100% (90%, 100%) | 93% (79%, 98%) |
| **HIV Negative** | 277 | 99% (96%, 100%) | 100% (98%, 100%) | 100% (98%, 100%) | 98% (96%, 99%) | 99% (97%, 100%) | 97% (94%, 99%) |
| **Underweight** | 51 | 98% (88%, 100%) | 100% (91%, 100%) | 100% (91%, 100%) | 98% (88%, 100%) | 100% (91%, 100%) | 90% (78%, 96%) |
| **Age < 5** | 185 | 98% (95%, 100%) | 99% (97%, 100%) | 100% (97%, 100%) | 98% (94%, 99%) | 100% (97%, 100%) | 95% (91%, 98%) |
| **Sensitivity by TB Case Criteria** |  |  |  |  |  |  |  |
| **Confirmed TB** | 153 | 33% (26%, 41%) | 12% (7%, 18%) | 12% (8%, 19%) | 14% (9%, 21%) | 15% (10%, 22%) | 37% (29%, 45%) |
| **Confirmed + Unconfirmed TB** | 310 | 19% (15%, 24%) | 6% (4%, 9%) | 6% (4%, 10%) | 9% (6%, 13%) | 7% (5%, 11%) | 21% (17%, 26%) |
| **Sensitivity by Subgroup,<sup>2</sup> % (95% CI)** |  |  |  |  |  |  |  |
| **HIV Positive** | 30 | 43% (26%, 62%) | 23% (11%, 43%) | 23% (11%, 43%) | 20% (8%, 39%) | 33% (18%, 53%) | 57% (38%, 74%) |
| **HIV Negative** | 116 | 31% (23%, 40%) | 9% (4%, 16%) | 9% (5%, 17%) | 13% (8%, 21%) | 10% (6%, 18%) | 32% (24%, 41%) |
| **Underweight** | 40 | 40% (25%, 57%) | 23% (11%, 39%) | 23% (11%, 39%) | 18% (8%, 33%) | 30% (17%, 47%) | 48% (32%, 64%) |
| **Age < 5** | 73 | 29% (19%, 41%) | 10% (4%, 19%) | 10% (4%, 19%) | 10% (4%, 19%) | 14% (7%, 24%) | 33% (23%, 45%) |

| **Sensitivity by Geographic Region,<sup>2</sup> % (95% CI)** |  |  |  |  |  |  |  |
| --- | --- | --- | --- | --- | --- | --- | --- |
| **The Gambia** | 30 | 40% (23%, 59%) | 30% (15%, 50%) | 33% (18%, 53%) | 20% (8%, 39%) | 33% (18%, 53%) | 43% (26%, 62%) |
| **South Africa** | 43 | 44% (29%, 60%) | 12% (4%, 26%) | 12% (4%, 26%) | 16% (7%, 31%) | 16% (7%, 31%) | 44% (29%, 60%) |
| **Uganda** | 23 | 13% (3%, 35%) | 4% (0%, 24%) | 9% (2%, 30%) | 17% (6%, 40%) | 13% (3%, 35%) | 35% (17%, 57%) |
| **Peru** | 34 | 18% (7%, 35%) | 0% (0%, 13%) | 0% (0%, 13%) | 0% (0%, 13%) | 3% (0%, 17%) | 18% (7%, 35%) |
| **India** | 23 | 48% (27%, 69%) | 13% (3%, 35%) | 9% (2%, 30%) | 22% (8%, 44%) | 9% (2%, 30%) | 43% (24%, 65%) |

AUC: Area under the ROC curve; CI: Confidence interval
1. Cut-Off selected for each assay based on ROC analysis of the paediatric sample set 2. Confirmed TB cases only

With the revised cut-off values, Ag85B had the highest sensitivity (33%, 95% CI 26-41) at 99% specificity (95% CI 97-100). This was similar sensitivity (p=0·47) and specificity (p=0·12) to LAM overall, with the same trends across subgroups and settings. Sensitivity continued to be higher in those with HIV and who were underweight, and lower in Uganda and Peru compared to the other study sites. Sensitivity again was reduced when Unconfirmed tuberculosis was included in the tuberculosis definition (Range 6-21%); 16 children (10%) with Unconfirmed tuberculosis had at least one *Mtb* protein or LAM detected.

### Combining Biomarker Measurements to Improve Sensitivity

**Figure 2** shows that urine concentrations of individual *Mtb* proteins and LAM are correlated (Pearson correlation coefficient = 0.72), but several samples had high levels of one of the antigens, but undetectable levels of the other. **Table 4** shows the most sensitive combinations of 2, 3, 4, 5 and 6 biomarkers. Combining LAM with Ag85B improved sensitivity to 45% for Confirmed TB, and 27% for clinically defined-cases (Confirmed or Unconfirmed tuberculosis), compared to 37% and 21%, respectively, for LAM alone (p<0·001 for both). Combining Ag85B, MPT32 and MPT64 with LAM increased the sensitivity according to the microbiological and composite reference standard to 50% and 30% (p<0·001 versus LAM alone), respectively, while maintaining a specificity of 94% (95% CI 91-96). Further increases in panel size provided no additional performance benefits.

**Figure 2.**
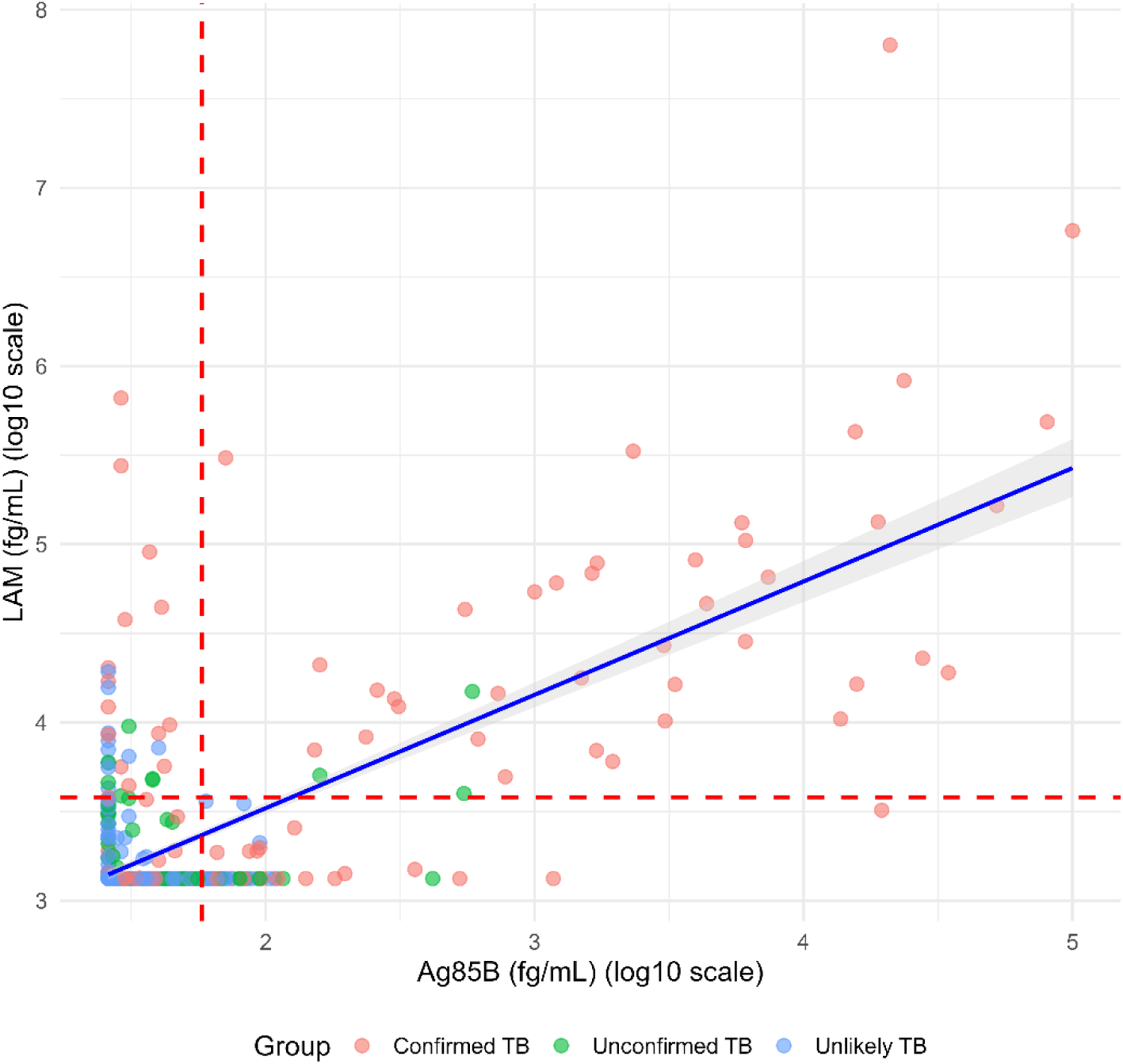
Correlation of the measured concentrations of LAM and Ag85B in the pediatric sample set. Points are colored based on tuberculosis status. The best linear fit with confidence limits is shown as a solid blue line with shading to indicate the 95% confidence limits. The Pearson correlation coefficient is 0.72. Assay cut-offs are indicated as dashed red lines.

**Table 4.** Performance of combining multiple urine *Mtb* antigens to detect childhood tuberculosis.

| Size | Panel | Specificity<br>% (95% CI) | Sensitivity % (95% CI) |  |  |  |  |  |
| --- | --- | --- | --- | --- | --- | --- | --- | --- |
|  |  | Unlikely TB | Confirmed +<br>Unconfirmed<br>TB | Confirmed TB Only |  |  |  |  |
|  |  |  |  | All | HIV- | HIV+ | Underweight | Age < 5 |
| **1** | **LAM** | 97% (94%, 98%) | 21% (17%, 26%) | 37% (29%, 45%) | 32% (24%, 41%) | 57% (38%, 74%) | 48% (32%, 64%) | 33% (23%, 45%) |
| **1** | **Ag85B** | 99% (97%, 100%) | 19% (15%, 24%) | 33% (26%, 41%) | 31% (23%, 40%) | 43% (26%, 62%) | 40% (25%, 57%) | 29% (19%, 41%) |
| **2** | **Ag85B, LAM** | 95% (92%, 97%) | 27% (22%, 32%) | 45% (37%, 53%) | 41% (32%, 50%) | 63% (44%, 79%) | 53% (36%, 68%) | 41% (30%, 53%) |
| **3** | **MPT32, Ag85B, LAM** | 94% (91%, 96%) | 29% (24%, 34%) | 48% (40%, 57%) | 45% (36%, 54%) | 63% (44%, 79%) | 58% (41%, 73%) | 45% (34%, 57%) |
| **4** | **MPT64, MPT32, Ag85B, LAM** | 94% (90%, 96%) | 30% (25%, 35%) | 50% (42%, 58%) | 47% (37%, 56%) | 63% (44%, 79%) | 60% (43%, 75%) | 47% (35%, 59%) |
| **5** | **ESAT6, MPT64, MPT32, Ag85B, LAM** | 93% (90%, 96%) | 30% (25%, 35%) | 50% (42%, 58%) | 47% (37%, 56%) | 63% (44%, 79%) | 60% (43%, 75%) | 47% (35%, 59%) |
| **6** | **ESAT6, CFP10, MPT64, MPT32, Ag85B, LAM** | 93% (90%, 96%) | 30% (25%, 35%) | 50% (42%, 58%) | 47% (37%, 56%) | 63% (44%, 79%) | 60% (43%, 75%) | 47% (35%, 59%) |

CI: Confidence interval

The improvements provided by the combined panels were most evident in children without HIV (Sensitivity 47%, 15% absolute difference vs. LAM alone, 95% CI 7-22, p< 0·001) than with HIV (Sensitivity 63%, 7% absolute difference vs. LAM alone, p=0·5) (**Table 4**). Specificity reduced slightly to 94% (95% CI 90-96) and 93% (95% CI 79-98), respectively. Among underweight children, the sensitivity increased to 60%, a 12% absolute increase over LAM alone, although not statistically significant (p=0·06). However, specificity dropped to 86% (95% CI 73-94) compared to 90% with LAM alone, though this was not statistically significant (p=0·5). Specificity for the four-biomarker panel remained high at 91% among children under 5 years.

## DISCUSSION

Urine-based tuberculosis testing has been limited to detection of LAM, primarily among individuals living with HIV. In this multicentre study, we demonstrate that multiple *Mtb* proteins can also be detected in paediatric urine using an ultrasensitive multiplex immunoassay, at the femtomolar level, expanding the spectrum of pathogen-derived biomarkers measurable in a non-invasive specimen. The individual protein assays showed consistently high specificity, and Ag85B demonstrated sensitivity comparable to LAM. Higher sensitivity was seen in children who were underweight as well as those with HIV. Importantly, combining multiple antigens (Ag85B, MPT64, MPT32 and LAM) improved detection compared with LAM alone, particularly among children without HIV, without a substantial loss of specificity. These findings highlight the feasibility and potential of multi-antigen urine detection to improve TB detection in children, and set benchmarks for additional diagnostic development.

Childhood tuberculosis is frequently paucibacillary, requiring pathogen-based assays to achieve extremely low limits of detection. The multiplex ECL platform enabled simultaneous quantification of five proteins at femtomolar concentrations, providing insight into antigen levels present in childhood TB disease. The observed paediatric-specific thresholds between 57 and 107 fg/mL are substantially below the detection limits achievable by conventional lateral flow assays, which typically operate in the pg–ng/mL range.^18^ These findings suggest that future point-of-care technologies will require substantial analytical advances, including signal amplification or analyte enrichment strategies. Although the ECL platform is not intended for routine clinical deployment in environments that are highly sensitive to infrastructure and cost considerations, it serves as an analytical reference system defining the sensitivity targets that next-generation urine diagnostics for TB must achieve.^13^

LAM has been the sole urinary biomarker available for tuberculosis diagnosis. However, we found that several *Mtb* proteins could also be detected in urine and support tuberculosis detection. LAM is a glycolipid that has structural heterogeneity depending on patient characteristics (i.e., HIV status, severity of tuberculosis disease) and sample type, and despite significant effort, the number of high quality antibodies that are available remains limited.^12,19^ We found that all proteins had high specificity regardless of age group that met the WHO target of ≥98%, an advantage over LAM assays where there have been false positives in young children due to cross reactivity with other bacteria and non-tuberculous mycobacteria when bagged samples are obtained.^7^ Among the five *Mtb* proteins evaluated, Ag85B showed the highest sensitivity and was similar to LAM, overall and across subgroups and clinical sites. This trend was seen whether we used an adult- or paediatric-specific cut-off, though specificities were higher with the paediatric cut-off. Ag85B is abundantly secreted by *Mtb* as part of the antigen 85 complex and has an important role in binding to alveolar macrophages and catalyzing cell wall assembly as mycolyl transferases. It has been a target for several vaccine candidates,^20^ but its role for tuberculosis diagnosis has been limited to sputum or serologic testing.^21^ The detection of Ag85B in urine represents a previously unreported observation and supports exploration of multi-antigen signatures rather than reliance on a single biomarker.

Although the other proteins (CFP-10, ESAT-6, MPT32, and MPT64) are all secreted by *Mtb* and associated with tuberculosis disease,^9,10^ they had lower sensitivity than Ag85B and LAM in urine samples. CFP-10 and ESAT-6 are used in interferon-gamma release assays, and there are plasma-based assays that have shown promise.^22^ MPT64 is a protein used to distinguish *Mtb* from other mycobacteria on culture samples, and MPT32 is a secreted protein that acts as adhesin to facilitate binding to epithelial cells.^23^ While past studies noted potential utility in adults with pulmonary and extrapulmonary tuberculosis,^9,10^ our results suggest that at least alone they have limited sensitivity to detect pulmonary tuberculosis disease in children.

Higher sensitivity among children living with HIV and those who were underweight likely reflects increased systemic antigen burden associated with disseminated or more advanced disease. Malnutrition and immunosuppression may both facilitate antigen release into circulation and subsequent renal excretion.^3,5^ Previous studies in Niger and Mozambique found that a third of children with severe acute malnutrition and tuberculosis symptoms were LAM positive.^24,25^ Children who are underweight may reflect both a risk factor for disseminated disease (i.e. immune dysregulation from malnutrition) as well as a consequence of more advanced or severe tuberculosis disease. Variation by HIV and nutritional status may also explain the differences in sensitivity by country, with lower proportions of both in Uganda and Peru associated with reduced biomarker performance. Importantly, specificity remained consistently high across all geographic regions, indicating that site-level variation in sensitivity was unlikely to be driven by assay noise or cross-reactivity.

Although urinary concentrations of *Mtb* proteins and LAM in urine were correlated, discordant detection patterns supported a multi-antigen approach. We found that a four-antigen signature (MPT64, MPT32, Ag85B, LAM) detected 50% of the Confirmed TB cases, while retaining high specificity. Although it did not meet the WHO TPP for diagnostic sensitivity (≥66%), it is important to recognize that it was higher than LAM alone and higher or comparable to other non-sputum assays that are used in clinical care, including Xpert testing of urine (0-14%),^26^ stool (56-61%),^27^ and nasopharyngeal aspirates (44%).^28^ There have been few studies on multi-antigen urine testing for TB;^29^ a rapid immunochromatography test has been developed for ESAT6, CFP10, and MPT64 that showed 68% sensitivity to detect pulmonary TB in adults, but only 33% specificity.^29^ Towards the ultimate application of the biomarkers in a diagnostic test, we prioritized cut-offs that retained high specificity albeit with a lower sensitivity. Among children with HIV, the sensitivity was closer to the target at 63% sensitivity, and is similar to the performance of sputum Xpert Ultra in children with HIV.^28^ The four-antigen signature particularly boosted sensitivity in children without HIV, with accuracies that were similar to LAM alone in children with HIV.^30^ Most children with TB do not have HIV, and so this has the potential to greatly expand urine testing for children.

This study represents, to our knowledge, the largest evaluation of urinary Mtb protein detection in children across diverse high-burden settings. Strengths include rigorous clinical classification, representation of key paediatric risk groups, and application of a highly sensitive multiplex ECL platform. However, several limitations should be acknowledged. Use of stored samples may have reduced detectable antigen concentrations despite standardized freezing procedures. The case– control sampling framework may overestimate specificity relative to real-world populations, and assay thresholds were optimized within related datasets, introducing potential optimism bias. External prospective validation will therefore be essential. Furthermore, although identified using an ultrasensitive analytical system, translation into affordable point-of-care formats remains a major developmental step.

In conclusion, we demonstrate that *Mtb* proteins are detectable in paediatric urine with high specificity and that multi-antigen signatures can augment sensitivity over LAM alone for the diagnosis of childhood tuberculosis. These findings increase our understanding of pathogen-derived urinary biomarkers, and establish the analytical benchmarks needed to develop a point-of-care, urine-based assay to support TB detection in children.

## GROUP AUTHORSHIP

COMBO Study co-authors include:

Ainebyona Aggrey^1^, Alfred Andama^1^, Cynthia Baard^2^, Martina Boakarie^3^, Madikoi Danso^3^, Marie P. Gomez^3^, Amita Gupta^4,5^, Yekiwe Hlombe^2^, Sheriff Kandeh^3^, Sarjo Koita^3^, Vidya Mave^5^, Francis S. Mendy^3^, Moses Nsereko^1^, Mary Mudiope^1^, Ezekiel Mupere^1^, Winnie Nabakka^1^, Jascent Nakafeero^1^, Juliet Namboowa^1^, Gertrude Nanyonga^1^, Margaretha Prins^2^, Binta Saidy^3^, Moorine Sekadde^1^, Hellen Aanyu Tukamuhebwa^1^, Lesley Workman^2^

1. World Alliance for Lung and Intensive Care Medicine in Uganda (WALIMU), Uganda
2. Department of Paediatrics and Child Health, South African Medical Research Council Unit on Child and Adolescent Health, University of Cape Town, Cape Town, South Africa
3. Medical Research Council Unit The Gambia at the London School of Hygiene and Tropical Medicine, The Gambia
4. School of Medicine, Division of Infectious Diseases, Johns Hopkins University, Baltimore, MD, USA
5. Center for Infectious Diseases in India, Johns Hopkins India, Pune, India

## Data Availability

De-identified individual participant data that underline the results reported in this article, with data dictionary, are available following publication by researchers who provide a methodologically sound proposal that is approved by the study investigators. Please contact the corresponding author to share a proposal; all requesters will also need to sign a data sharing agreement.

## ACKNOWLEDGEMENTS

The authors thank the physicians, nurses, and staff at our multiple clinical sites who provided care for the children in this study. Additionally, the authors are grateful to the children with TB disease and their parents who were willing to participate in the study and help contribute meaningful data that may help future patients with the same illness. BEI resources (National Institute of Allergy and Infectious Diseases) provided reagents (Mycobacterium tuberculosis, Strain H37Rv, Purified Lipoarabinomannan (LAM), NR-14848, and Culture Filtrate Proteins, NR-14825.)

This research has been conducted using the FIND Specimen Bank lipoarabinomannan (LAM) antibodies and associated clinical samples.

## POTENTIAL CONFLICTS OF INTEREST

SM, MW, JW and GBS are all employees, and JW has an ownership interest, at Meso Scale Diagnostics, LLC. MSD and the funding agencies did not have any role in the decision to publish.

## FUNDING

This study was supported by the National Institute of Allergy and Infectious Diseases, National Institutes of Health (R01AI152161 to AC and JE, U01AI152087 to AC, U19AI109755 to MF). The data from India in this manuscript were collected as part of the Regional Prospective Observational Research for Tuberculosis (RePORT) India Consortium, funded in whole or in part with Federal funds from the Government of India’s (GOI) Department of Biotechnology (DBT), the Indian Council of Medical Research (ICMR), the United States NIH, NIAID, Office of AIDS Research (OAR), and distributed in part by CRDF Global; and the RePORT International Coordinating Center (RICC supplemental grant: DAA3-18-64080-1).

## Notes

### Author Declarations

The Ethics Research Committee of the Mulago Hospital gave ethical approval for this work. The Joint Ethics Committee of the Gambian Government and MRC gave ethical approval for this work. The Research Ethics Committee of the London School of Hygiene and Tropical Medicine gave ethical approval for this work. The Institutional Ethics Committee of the Research of National Institute of Health - Peru gave ethical approval for this work. The Human Research Ethics Committee of the Faculty of Health Sciences, University of Cape Town, gave ethical approval for this work. The Ethics Committee of the BJ Government Medical College gave ethical approval for this work. The Institutional Review Board (IRB) of Johns Hopkins University (JHU) gave ethical approval for this work. The IRB of the University of California, San Francisco (UCSF) gave ethical approval for this work.

