## Supplemental Information for "The accuracy of urine-based mycobacterial antigens to detect childhood tuberculosis using an ultrasensitive immunoassay"

**SUPPLEMENTAL MATERIAL**

**Table of Contents**

***Supplemental Methods*** **2**

***Supplemental Table 1. Classification performance of MTB Protein 5-Plex and LAM Assays with the adult cohort*** **7**

***Supplemental Table 2. Classification performance of Mtb Protein 5-Plex and LAM Assays with the paediatric cohort using cut-off values optimized for the adult cohort*** **8**

***Supplemental Figure 1. Representative calibration curves for the Mtb protein and LAM assays*** **10**

***Supplemental Figure 2. Measured concentrations of Mtb biomarkers in the adult urine sample set*** **11**

***Supplemental Figure 3. ROC curves for the Mtb protein and LAM assays showing the ability of the assays to classify (a) adult samples as tuberculosis positive or negative and (b) pediatric samples as Confirmed tuberculosis or Unlikely tuberculosis*** **12**

### Supplemental Methods

#### Paediatric Clinical Procedures

Symptoms suggestive of tuberculosis included cough of any duration and at least one of the following: weight loss or poor weight gain, fever lasting more than one week, lethargy or reduced playfulness, or known tuberculosis contact. We excluded children who had received more than three doses of anti-tuberculous treatment, those who had completed tuberculosis treatment in the preceding 12 months, or those who were unable to provide respiratory samples for microbiological testing. Written informed consent was obtained from parents or guardians, with assent from older children as appropriate. Ethical approval was obtained from the Mulago Hospital Ethics Research Committee, Gambian Government and MRC Joint Ethics Committee, London School of Hygiene and Tropical Medicine Research Ethics Committee, Institutional Ethics Committee for Research of National Institute of Health - Peru, Human Research Ethics Committee of the Faculty of Health Sciences, University of Cape Town, Ethics Committee at BJ Government Medical College, Johns Hopkins University (JHU) Institutional Review Board (IRB), and the University of California, San Francisco (UCSF) IRB.

Clinical histories and physical examinations were obtained for all children. All participants were tested for HIV infection, and chest radiographs were obtained. Induced or expectorated sputum, gastric aspirates, or nasopharyngeal aspirates were obtained for smear microscopy, Xpert MTB/RIF or Xpert Ultra, and mycobacterial culture. Urine samples were collected during the same visit as respiratory specimens, following standardised techniques. Children unable to void into a collection cup had a bagged specimen obtained. Samples were aliquoted and stored at –80°C until testing. The decision to initiate tuberculosis treatment was made by the treating clinicians, following national guidelines and based on clinical evaluation and available test results. All participants had follow-up at 2-3 months to assess clinical status.

#### ***Mtb*** **Protein Assay Reagents**

Five Mtb protein targets were expressed recombinantly with a C-terminal His6 purification tag from an E. coli expression system for use as assay calibrators and as immunogens for antibody development. The proteins (with long name, gene ID and TB protein ID) were Ag85B (Antigen 85b, fbpB, Rv1886c), CFP-10 (Culture Filtrate Protein 10 kDa, esxB, Rv3874), ESAT-6 (Early Secretory Antigenic Target 6 kDa, esxA, Rv3875), MPT32 (MPT32, apa, Rv1860) and MPT64 (MPT64, mpt64, Rv1980c). The expressed protein sequences were based on the H37Rv reference Mtb strain (NCBI reference sequence NC_000962.3) as annotated in EPFL Mycobrowser (https://mycobrowser.epfl.ch/). All proteins were purified by nickel affinity chromatography and additionally by anion exchange and/or size exclusion chromatography to greater than 95% purity. Purified LAM and culture filtrate from the Mtb H37Rv strain were obtained from BEI Resources (catalog numbers NR-14848 and NR-14825, respectively).

Monoclonal antibodies against the five protein targets were generated by immunising mice with recombinant proteins and screening antibodies using conventional hybridoma methods. Antibodies with binding activity were produced at pilot scale and labeled with biotin and the ECL SULFO-TAG label from MSD to be tested as capture and detection antibodies, respectively, in an ECL sandwich immunoassay format. Optimal antibody pairs were selected based on sensitivity in detecting both purified recombinant protein and native protein in Mtb cell culture filtrates. For ESAT-6 and CFP-10, additional improvements in sensitivity were obtained by using a capture antibody consisting of a 1:1 mixture of two monoclonal antibodies recognising two different epitopes. Antibodies selected for use in the final multiplexed assay format (described below) were sequenced, produced recombinantly in a mouse IgG1 framework by transient expression in CHO cells, and purified by Protein A affinity chromatography and size exclusion chromatography. Antibodies against LAM – clones A194-01, S4-20 and FIND28 - were originally developed at Rutgers, Otsuka, and the Foundation for Innovative New Diagnostics (FIND),[1] respectively, and were provided by FIND.

***Mtb* Protein 5-Plex and LAM Assay Protocol**

Briefly, the Mtb protein 5-plex procedure involved five incubation steps. The plates were washed before and after each step (3 x 300 uL of PBS per well), and incubations were carried out at 27°C with shaking in a heated shaker. First, 50 uL of a solution containing a mixture of the capture antibodies for the five assays was added to each well and incubated for 1 hour with shaking. Each of the capture antibodies in the mixture was labeled with biotin and coupled through the biotin a “linker” that uniquely binds one of the spots, allowing for self-assembly of the capture antibodies into an array. Second, 30 µL of Blocking Solution and 20 µL of sample were added to each well, incubating for 90 minutes with shaking to capture analyte from the samples. Third, 50 µL of a solution of the five detection antibodies (labeled with the S-PLEX TURBO-BOOST® label) in Antibody Diluent was added and incubated for 1 hour to complete the sandwich complex. Fourth, the S-PLEX signal enhancement step was initiated by adding 50 µL of S-PLEX Enhance Solution and incubating 30 minutes. Fifth, the signal enhancement step was completed by adding 50 µL of S-PLEX TURBO-TAG Detection Solution and incubating

60 minutes. MSD Read Buffer B was then added to each well and ECL signals were measured using a MESO SECTOR S 600MM or MESO QuickPlex SQ 120MM ECL plate reader.

The LAM assay was run as previously reported[1] with two differences: (i) the heat pre-treatment step used in the original protocol was found to not provide any performance benefit with urine samples, so samples were run undiluted without any pre-treatment, and (ii) the original commercial purified LAM used to calibrate the assay was no longer available and was replaced with purified LAM from BEI resources, potentially leading to small differences in absolute quantitation by the assay.

**Assay Calibration**

Calibration data for assay on each plate was fit, using MSD Methodical Mind® software, to a four-parameter-logistic (4PL) model with $1/Y^2$ weighting. Concentrations for unknown samples and controls were calculating by back-fitting to the 4PL model and averaging across replicates. Calculated concentrations below one half of the assay limit of detection (LOD / 2) were assigned the value of LOD/2 for statistical analysis and plotting. Calculated concentrations above the highest calibrator dilution (the “top-of-curve” or TOC) were assigned the value of TOC.

The LOD and LLOQ for each assay was determined by running six assay plates with duplicate calibration curves, 20 blanks per plate, and a set of LLOQ samples run in triplicate on each plate. The LLOQ samples were designed to cover a range of concentrations near the LOD, and were prepared by diluting the calibration standard. The LOD was calculated as the concentration that gives a signal 2.5 standard deviations (SDs) above the average blank signal. The presented LOD value is the median of the

LOD calculated on each of the 6 individual plates. The presented LLOQ is the lowest concentration that provided an average calculated concentration between 80% and 120% of the expected value, and a coefficient of variation (CV) less than 25%.

### Supplemental Table 1. Classification performance of MTB Protein 5-Plex and LAM Assays with the adult cohort

| | | N | Mtb Protein 5-Plex | | | | | |
|---|---|---|---|---|---|---|---|---|
| | | | **Ag85B** | **CFP10** | **ESAT6** | **MPT32** | **MPT64** | **LAM** |
| **Cut-Off (fg/mL)**[1] | | | 57 | 107 | 63 | 76 | 87 | 3,810 |
| **AUC** | | | 0·78 | 0·79 | 0·85 | 0·6 | 0·79 | 0·9 |
| **Specificity % (95% CI)** | | 30 | 97% (81%, 100%) | 97% (81%, 100%) | 97% (81%, 100%) | 97% (81%, 100%) | 87% (68%, 96%) | 93% (76%, 99%) |
| **Sensitivity % (95% CI)** | **All** | 30 | 63% (44%, 79%) | 43% (26%, 62%) | 57% (38%, 74%) | 23% (11%, 43%) | 63% (44%, 79%) | 87% (68%, 96%) |
| | **HIV-** | 15 | 40% (17%, 67%) | 33% (13%, 61%) | 47% (22%, 73%) | 7% (0%, 34%) | 47% (22%, 73%) | 87% (58%, 98%) |
| | **HIV+** | 15 | 87% (58%, 98%) | 53% (27%, 78%) | 67% (39%, 87%) | 40% (17%, 67%) | 80% (51%, 95%) | 87% (58%, 98%) |

AUC: Area under the ROC curve; CI: Confidence interval

1. Cut-Off is the preliminary cut-off selected for each assay based on ROC analysis of the adult sample set

**Supplemental Table 2. Classification performance of *Mtb* Protein 5-Plex and LAM Assays with the paediatric cohort using cut-off values optimized for the adult cohort**

| | | *Mtb* Protein 5-Plex | | | | | |
|---|---|---|---|---|---|---|---|
| | **N** | **Ag85B** | **CFP10** | **ESAT6** | **MPT32** | **MPT64** | **LAM** |
| **Cut-Off (fg/mL)**[1] | NA | 57 | 107 | 63 | 76 | 87 | 3810 |
| **AUC** | NA | 0·72 | 0·52 | 0·58 | 0·6 | 0·62 | 0·7 |
| **Specificity, % (95% CI)** | | | | | | | |
| **Unlikely TB** | 320 | 94% (90%, 96%) | 92% (88%, 94%) | 95% (91%, 97%) | 98% (96%, 99%) | 82% (77%, 86%) | 97% (94%, 98%) |
| **HIV Positive** | 42 | 95% (83%, 99%) | 95% (83%, 99%) | 93% (79%, 98%) | 98% (86%, 100%) | 76% (60%, 87%) | 93% (79%, 98%) |
| **HIV Negative** | 277 | 94% (90%, 96%) | 91% (87%, 94%) | 95% (91%, 97%) | 98% (96%, 99%) | 83% (78%, 87%) | 97% (95%, 99%) |
| **Underweight** | 51 | 86% (73%, 94%) | 94% (83%, 98%) | 94% (83%, 98%) | 96% (85%, 99%) | 78% (64%, 88%) | 92% (80%, 97%) |
| **Age < 5** | 185 | 92% (87%, 95%) | 91% (86%, 95%) | 94% (89%, 97%) | 97% (93%, 99%) | 79% (72%, 84%) | 96% (91%, 98%) |
| **Sensitivity by TB Case Criteria, % (95% CI)** | | | | | | | |
| **Confirmed TB** | 153 | 39% (31%, 47%) | 17% (12%, 24%) | 16% (11%, 23%) | 14% (9%, 21%) | 33% (25%, 41%) | 35% (27%, 43%) |
| **Confirmed + Unconfirmed TB** | 310 | 23% (18%, 28%) | 10% (7%, 14%) | 9% (6%, 13%) | 9% (6%, 13%) | 23% (19%, 28%) | 20% (16%, 25%) |
| **Sensitivity by Subgroup,[2] % (95% CI)** | | | | | | | |
| **HIV Positive** | 30 | 50% (33%, 67%) | 23% (11%, 43%) | 30% (15%, 50%) | 20% (8%, 39%) | 50% (33%, 67%) | 50% (33%, 67%) |
| **HIV Negative** | 116 | 36% (28%, 46%) | 16% (10%, 24%) | 13% (8%, 21%) | 13% (8%, 21%) | 29% (21%, 39%) | 31% (23%, 40%) |
| **Underweight** | 40 | 48% (32%, 64%) | 23% (11%, 39%) | 30% (17%, 47%) | 18% (8%, 33%) | 48% (32%, 64%) | 45% (30%, 61%) |
| **Age < 5** | 73 | 33% (23%, 45%) | 11% (5%, 21%) | 12% (6%, 23%) | 10% (4%, 19%) | 30% (20%, 42%) | 30% (20%, 42%) |
| **Sensitivity by Geographic Region,[2] % (95% CI)** | | | | | | | |
| **The Gambia** | 30 | 47% (29%, 65%) | 33% (18%, 53%) | 40% (23%, 59%) | 20% (8%, 39%) | 63% (44%, 79%) | 40% (23%, 59%) |
| **South Africa** | 43 | 49% (34%, 64%) | 12% (4%, 26%) | 12% (4%, 26%) | 16% (7%, 31%) | 30% (18%, 46%) | 42% (27%, 58%) |
| **Uganda** | 23 | 17% (6%, 40%) | 9% (2%, 30%) | 9% (2%, 30%) | 17% (6%, 40%) | 26% (11%, 49%) | 30% (14%, 53%) |
| **Peru** | 34 | 21% (9%, 38%) | 6% (1%, 21%) | 3% (0%, 17%) | 0% (0%, 13%) | 12% (4%, 28%) | 18% (7%, 35%) |
| **India** | 23 | 57% (35%, 76%) | 30% (14%, 53%) | 22% (8%, 44%) | 22% (8%, 44%) | 35% (17%, 57%) | 43% (24%, 65%) |

AUC: Area under the ROC curve; CI: Confidence interval

1. Cut-Off is the preliminary cut-off selected for each assay based on ROC analysis of the adult sample set
2. Confirmed TB cases only

**Supplementary Figure 1. Representative calibration curves for the *Mtb* protein and LAM assays.** Points show the assigned concentrations and measured ECL signals for the individual calibration samples. The calculated 4PL fit is shown as a solid line. The vertical dashed lines indicate the LOD (blue), LLOQ (green) and TOC (red) concentrations.

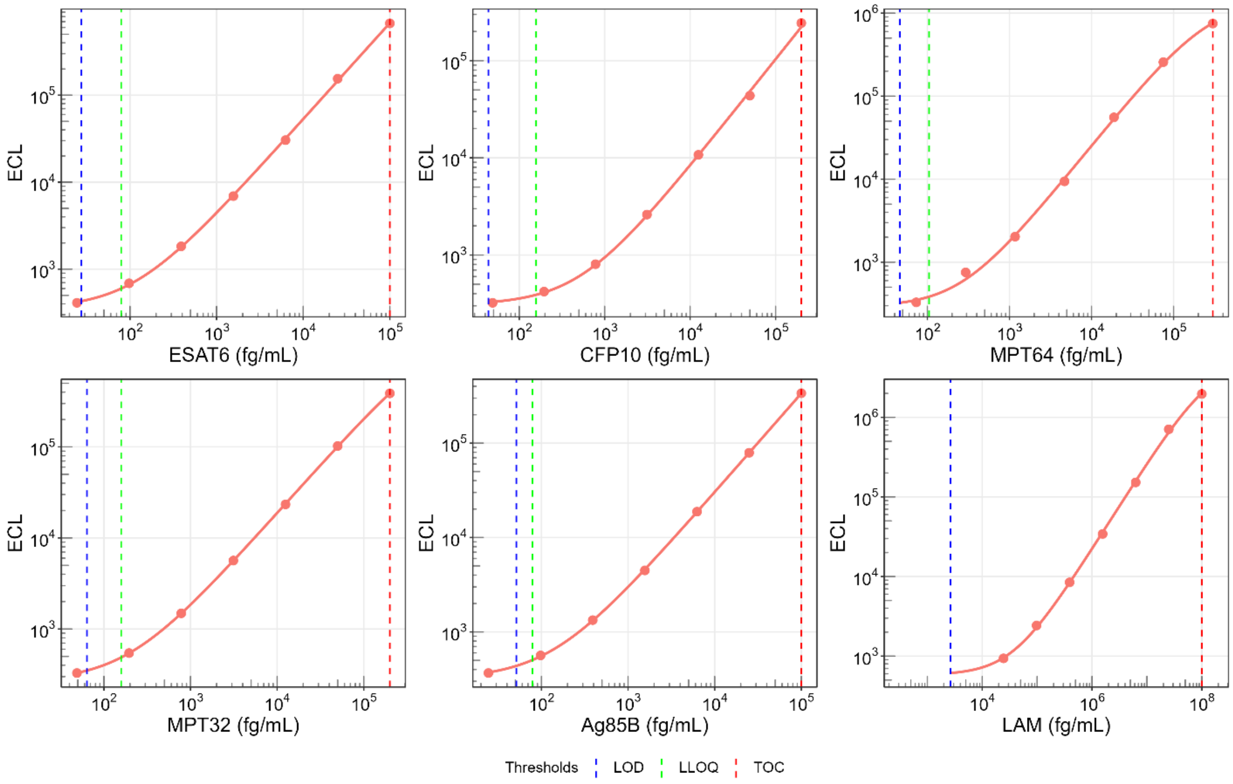

**Supplementary Figure 2. Measured concentrations of *Mtb* biomarkers in the adult urine sample set.** Points are colored to indicate country, and shaped based on HIV status. The dashed horizontal lines indicate the preliminary cut-off values based on ROC curve analysis of the adult samples (green).

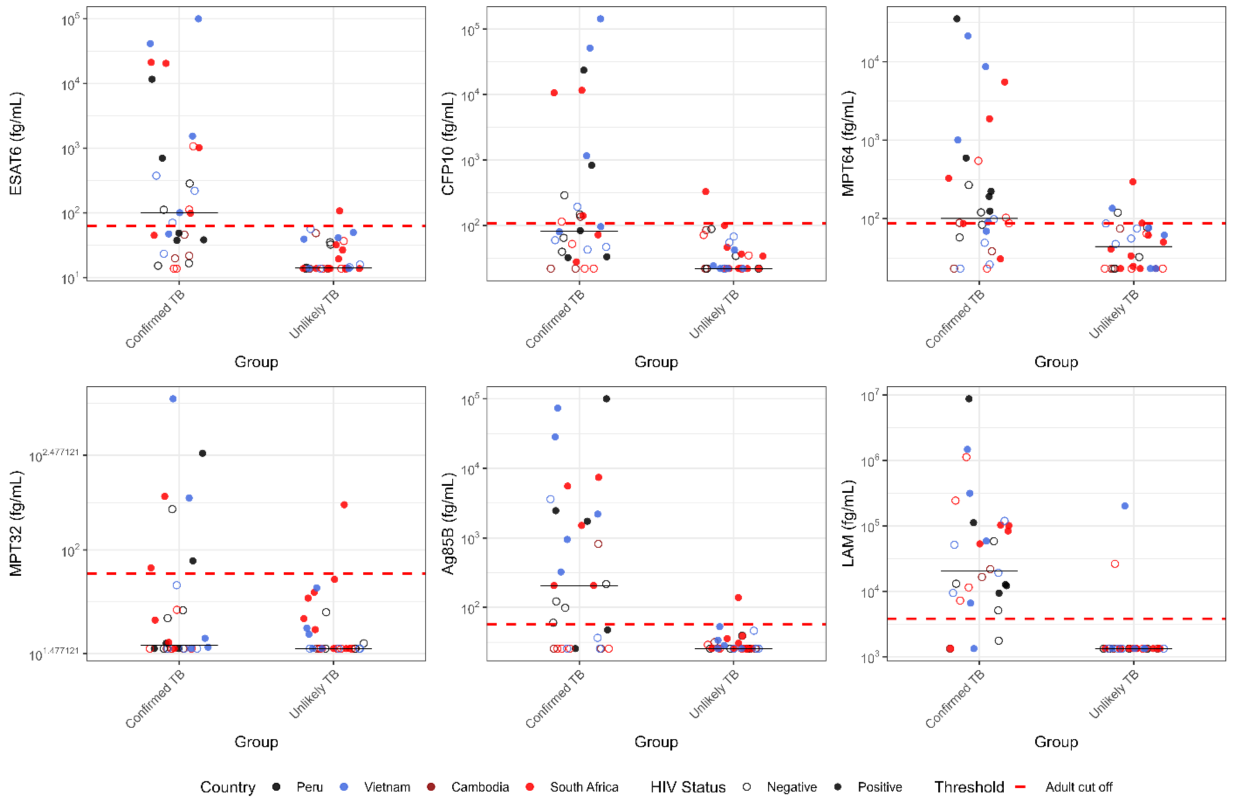

**Supplementary Figure 3. ROC curves for the *Mtb* protein and LAM assays showing the ability of the assays to classify (a) adult samples as tuberculosis positive or negative and (b) pediatric samples as Confirmed tuberculosis or Unlikely tuberculosis.** The preliminary cut-off value selected based on ROC analysis of the adult samples is indicated with a blue dot. The re-optimized cut-off value selected based on the paediatric data set is indicated with a red dot.

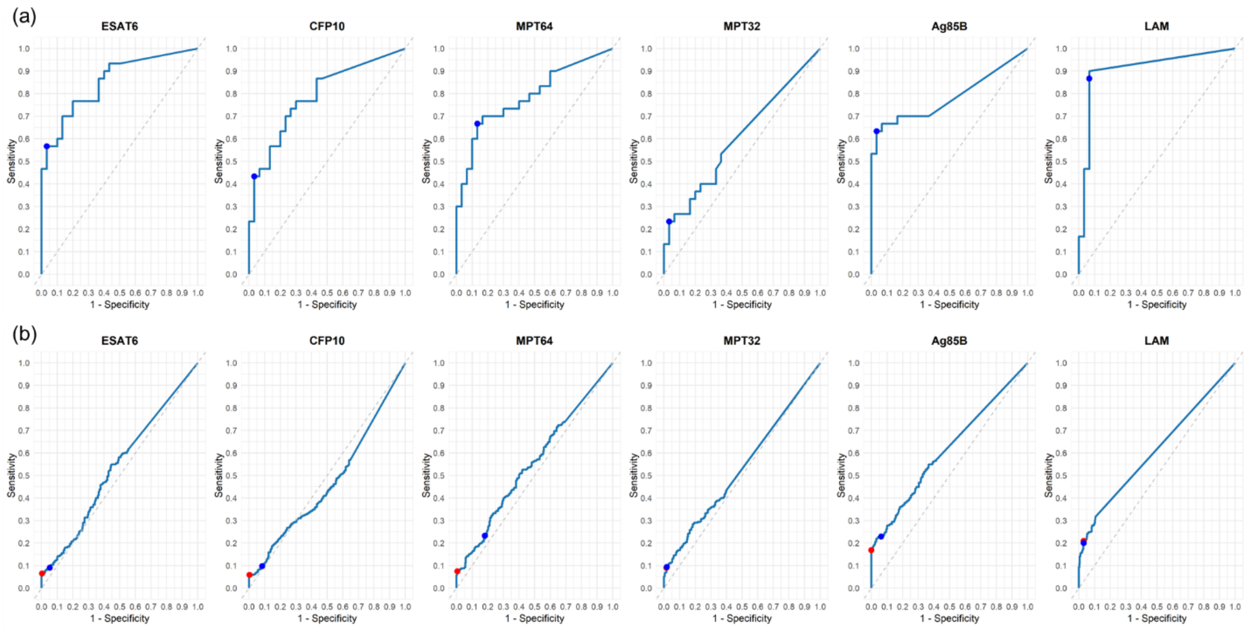
